# Is family social exclusion associated with child psychomotor and social development delay in southeastern Brazil?

**DOI:** 10.1101/2022.05.26.22275658

**Authors:** Clariana Vitória Ramos de Oliveira, Cláudia Nery Teixeira Palombo, Joshua Jeong, Katherine Maria Solís Cordero, Elizabeth Fujimori

**Affiliations:** School of Nursing, University of Nevada Las Vegas; School of Nursing, Federal University of Bahia, Salvador, Bahia, Brazil; Department of Global Health and Population, Harvard T.H. Chan School of Public Health, Boston, Massachusetts, United States of America; School of Nursing, University of Costa Rica, San Jose, Costa Rica; Department of Nursing Public Health, School of Nursing, University of São Paulo, São Paulo, Brazil

**Keywords:** Child development, Cross-Sectional Study, Social Disadvantage, Social Deprivation, Developing Countries

## Abstract

The early years of life are a crucial period for shaping child development. Children from socially excluded families may have less opportunity to attain their full development. The objective was to identify if family social exclusion is associated with child psychomotor and social development delay in Southeastern Brazil. A cross-sectional study was conducted using data from a sample of 348 children under three years. We measured psychomotor and social development delays. An index was used to evaluate social exclusion. The prevalence of child psychomotor and social developmental delay was 27.6% and 17.2%, respectively. Children in the most social excluded group were more likely to have delayed psychomotor development (OR = 3.4; 95% CI = 1.14; 10.55) and social developmental delay (OR = 3.9; 95% CI = 1.05; 9.02) than children in the least social excluded group. The results confirm the association between family social exclusion and child development delay.

## Introduction

The early years of life are crucial for shaping children’s developmental potential across the life course. However, an estimated 250 million children under five years in low- and middle-income countries are at risk of not reaching their full learning, social, and emotional development potential, which has a negative impact on their future [1]. The family’s low socioeconomic status position has been associated with a negative impact on the neurocognitive domains including language and executive functioning [2].

Recognition of the importance of early childhood development (ECD) can be evidenced by the growth of countries that have adopted multi-sectoral policies to promote ECD and their inclusion among the 2015-2030 Sustainable Development Goals [3]. Over the past three decades, Brazil has shown a substantial improvement in child health indicators with improvements in child survival, however this occurs unequally within country and among the income quintiles [4-5]. Therefore, it is still necessary to overcome the inequalities of the most vulnerable groups to ensure that children achieve healthy and holistic development. In early childhood, biological and social determinants directly affect children’s brain, cognitive, social, and emotional development [6]. More specifically, poverty, lack of stimulation, excessive stress, poor caregiver-child relationships, and the surrounding social environment have been identified as significant risk factors for child development [6-7]. Social inequality and adverse early childhood experiences have long-term physiological and epigenetic impact on brain development and cognition [8].

Studies conducted in different regions of the world show that ECD is closely related to the family’s socioeconomic situation. A study from India, Indonesia, Peru, and Senegal revealed socioeconomic gradients of early child development and specifically found that children under two years from the wealthiest households had higher developmental scores and better growth than children from the poorest households [9]. Research in Iran has found that home affordance, parental education, house space, availability of toys, and attending daycare were associated with children’s motor development [10].

However, a growing body of research indicates that economic poverty alone does not explain the large disparities in maternal and child health care between rich and poor families. Consequently, it is necessary to develop new methodologies and guidelines that can address social inequality and socioeconomic positions that can link these measures with health [11]. Without specific measures to address the family social roots, social inequality in health cannot be addressed [12]. A systematic review found that social excluded families were affected by greater health problems than privileged social families [13] and new approaches to analyze wealth inequalities have been proposed, despite being widely studied [14]. Family social exclusion has been considered by several Latin American researchers [15-17] as specific indicator to understand the health-disease process and analyze social determinants.

A study from Australia observed that social exclusion – based on various indicators including poor accommodation, unemployment of mother, no access to a car, difficult financial situation, marital status, low education of mother, small social support network, poor self-reported mother’s health, and unplanned pregnancy – had a negative impact on maternal and child health [18]. A study from South Korea found that maternal job strain during the pregnancy is a determinant of infant neurodevelopment delay at 6 and 12 months [19]. In China, a study observed that social inequality was associated with poor neurodevelopment for children under three years in a poor rural area [20]. In Peru, a study highlighted that children between 0 to 5 years in rural areas whose mothers had low schooling and lived in households with unsatisfied basic needs had lower developmental scores [21]. Researchers in Colombia showed that children that lived in poor areas had more development problems than children that lived in rich areas [22]. A cohort study in three different cities in Brazil showed a stagnation on reducing rates of child mortality that could be associated with economic decline and increased poverty [5].

Besides poverty, the magnitude of social exclusion indicators in Brazil have not been fully elucidated. Thus, this study is based on the operational model of Trapé (2011), that developed a social exclusion index that accesses differences in social exclusion between families, taking into account different variables, with more precise evaluation for this population. This new index allowed us to investigate the relationship between social exclusion and early child psychomotor and social development in Southeastern Brazil.

## Materials and methods

### Study design and sample

This cross-sectional study analyzed data from a broader study entitled “Effect of nutritional counseling from Integrated Management of Childhood Illness strategy on eating habits, nutritional status and child development” [23]. The study was conducted in a sample of 348 children under three years of age, enrolled at 12 primary health care centers of a municipality with 48,000 inhabitants, located in São Paulo State, in Southeastern Brazil. The municipality is located 70 km west of the State’s capital, which is Brazil’s financial center, and the Southeastern region is one of the most developed, richest, yet with higher social inequality in the country.

For the broader study, the sample size was estimated based on the number of children under three years of age registered at all 12 primary care centers (n=3904), 50% prevalence rate of children with inadequate food practices, 95% confidence level and 5% margin of error, which indicating a necessary sample size of 350 children. Inclusion criteria were: child under 3 years of age who resided with their biological mother in the study municipality. In the case of more than one child younger than three years, the youngest child was selected for participation. We excluded non-biological children, twins, children with health problems such as metabolic syndrome, genetic and/or neurological problems, and children with a previous diagnosis of sickle cell anemia. Ethical approval was granted by the Research Ethics Committee of the University of São Paulo, Brazil (Process No. 193468). A informed consent was obtained from all participants and parent/guardian of each participant under 18 years of age.

### Data collection and variables

#### Outcome

ECD was measured using the “Developmental Surveillance Instrument” [24] which was developed by the Ministry of Health in Brazil, based on adaptations of the Denver-II [25] and Gesell [26]. This tool is organized into 11 age groups, which correspond to different developmental stages recommended for consultations of children between 0 and 6 years of age [24, 27]. For each age group, there are four indicators related to the domains maturative, psychomotor, social and cognitive to follow the process of child development [24]. Although it does not provide a diagnosis of child development, this instrument serves as a guide for primary care practitioners to monitor whether children are able or not able to attain basic developmental milestones (e.g. the child lying face down holds the head for 1 second; Recognize when someone is talking to him) and identify children at risk of potential developmental delays. For this study, we focused on children’s psychomotor (refers to the gradual process of refinement and integration of biomechanical skills and principles of movement, so that the result is consistent and effective motor behavior) and social development (that refers to the social interaction of the baby to the mother and others) [24].

Children’s ability to attain milestones was observed by the interviewers (two nurses, one master student and one doctoral student), and when necessary, the mother was asked about the acquisition of a certain skill. We followed the recommendation of the Ministry of Health [24] to identify children with healthy development according to the child’s age (children who attained 90% of the developmental milestones in the domain for the age group) or children presenting developmental delay (children who attained less than 90% of reached milestones development in the domain for the age group).

#### Exposure

The present study used a pre-tested instrument proposed by Trapé (2011) in order to operationalize an index that could measure family social exclusion in Brazil. This index was constructed based upon theories and methods from the field of social determinants of health [28], including the role of work as it relates to family life (consumption based on the capitalist accumulation process). The families were interviewed regarding social inclusion in society. Trapé (2011) initially used a set of 34 empirical variables to represent the concept of social inclusion of families who lived in different social spaces, from a representative sample from a municipality in the Metropolitan Region of São Paulo. The data were analyzed using multivariate statistical analysis and factor analysis to select those variables statistically significant and to establish the cutoff points for the composition of the four social groups (GS) [15]. As a last step, a discriminant linear analysis was performed, which again tested the variables to assess the degree of importance of each one, which generated equations for the construction for the social classification of families. The final variables that better represent social exclusion are presented in the Fig 1. Three family social groups were constructed: least social excluded, moderately excluded, and most excluded [15].

**Fig 1.**
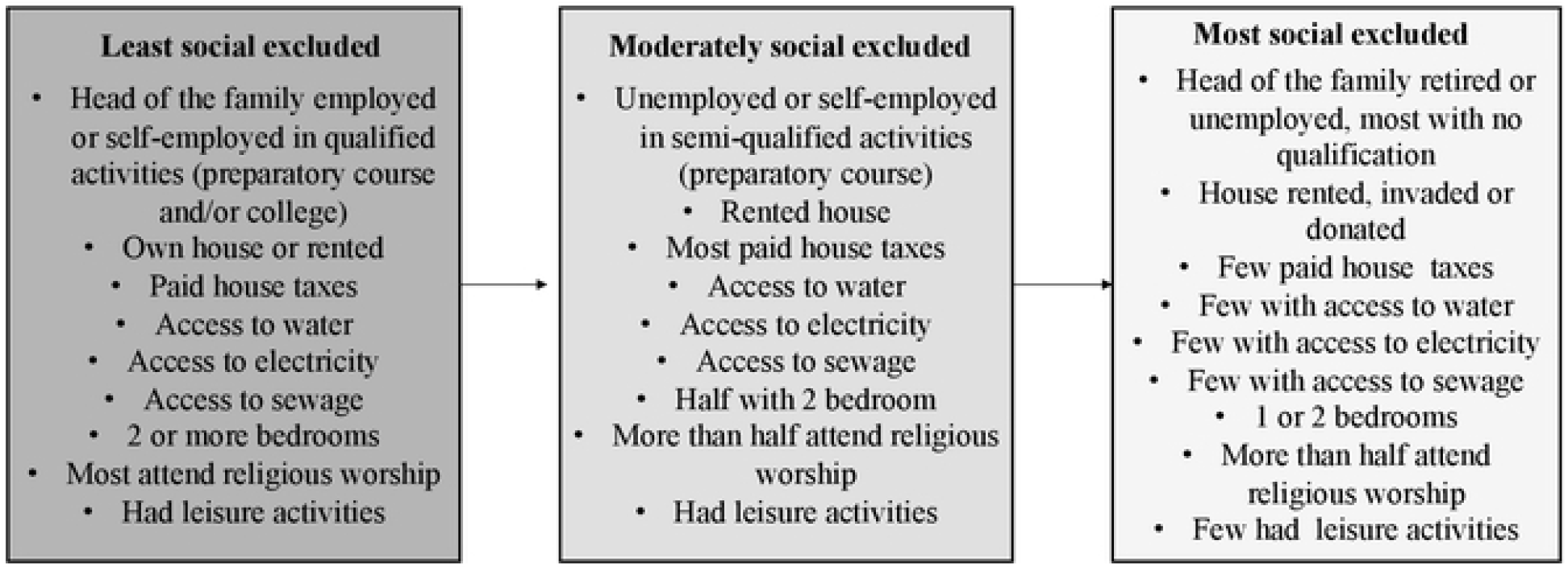
Family social groups categories.

#### Covariates

Drawing on the available evidence of children presenting developmental delays on the literature, four types of covariates were included in the analysis: (i) whether the family was a beneficiary of “Bolsa Família”, the conditional cash transfer program in Brazil (no or yes), (ii) family sociodemographic characteristics: maternal age (≤ 20, 20-29, and ≥ 30 years), maternal and paternal education (<8 and ≥8 of former schooling), number of siblings (1, 2 or ≥3; (iii) clinical and obstetric history: number of antenatal care visits (< 6 or ≥ 6), type of delivery (vaginal or cesarean section); (iv) child characteristics: child age; birth weight - collected from the child health booklet (<2500g and ≥2500g), hospitalization in the last 12 months (yes or no), breastfeeding (yes or no), and whether child attended day care (yes or no).

### Statistical analysis

To determine the relationship between exposures and ECD domain outcomes we used one-way ANOVA and χ2 tests. The association between children presenting developmental delays and its possible determinants was initially examined using univariate analysis and then following with hierarchical multiple logistic regressions using STATA software version 14.1. The hierarchical multiple regression blocks were tested to estimate the association between the exposure (family social groups) upon including additional relevant covariates in each model. We considered two dependent variables: children with psychomotor and social developmental delays. In the dimension of societal structural processes, we considered in block 1 the relationship between family social group and child development outcomes. In the dimension of processes in the child’s immediate environment, we added to the block 2 maternal characteristics: mother age, maternal occupation, number of siblings and Bolsa Família. Moreover, in block 3 we added obstetric and prenatal characteristic: number of antenatal care appointments and type of delivery. In the dimension of the child’s individual processes, we added child characteristics such as child age, gender, birth weight, hospitalization in the last 12 months, breastfeeding, and day care attendance. The magnitude of the association between dependent and independent variables was estimated by the odds ratio (OR) and their respective 95% confidence intervals and p-value <0.05. The hierarchical logistic regressions were performed according to the theoretical model defined a priori (Fig 2).

**Fig 2.**
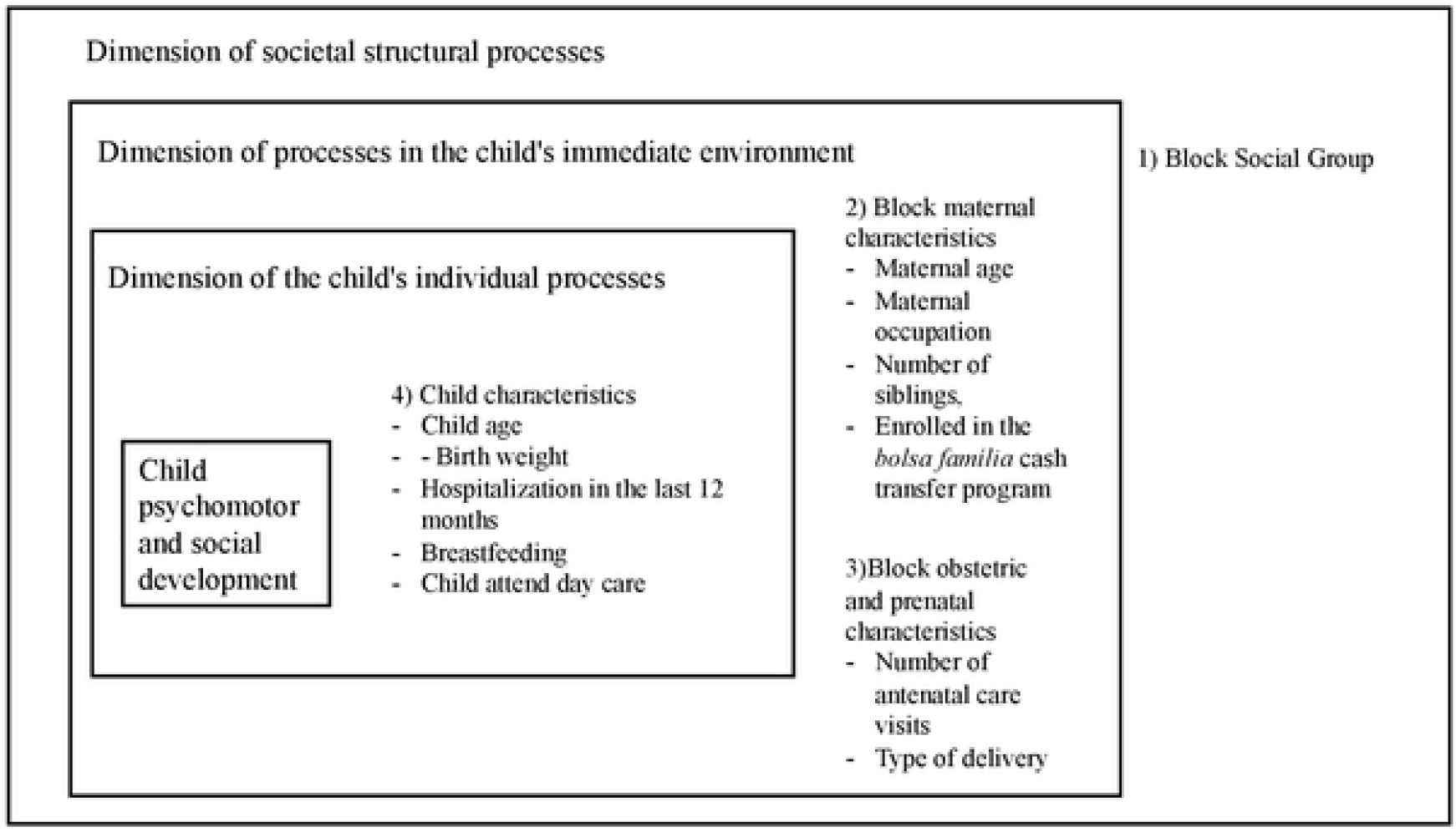
Conceptual model for the analysis of the child development.

## Results

The sample comprised of 358 children under 3 years of age. The average child age was 27.4 months and 54.6% were female. In the sample, 27.6% of children presented with psychomotor developmental delay and 17.2% children presented with social developmental delays. Study population characteristics are presented in Table 1.

**Table 1.** Family social groups, family and child characteristics, according to psychomotor and social development in children under 3 years old (n = 358). Municipality of São Paulo State, Brazil, 2013.

| Variables | Total<br>N (%) | Psychomotor<br>development |  | P-value | Social<br>development |  | P-value |
| --- | --- | --- | --- | --- | --- | --- | --- |
|  |  | Delay<br>N (%) | Adequate<br>N (%) |  | Delay<br>N (%) | Adequate<br>N (%) |  |
| Family social groups |  |  |  | 0.02 |  |  | 0.04 |
| Least social excluded | 69(19.8) | 11(11.5) | 58(23.0) |  | 8(13.3) | 61(21.2) |  |
| Moderately social excluded | 142(40.8) | 39(40.6) | 103(40.9) |  | 20(33.3) | 122(42.4) |  |
| Most social excluded | 137(39.4) | 46(47.9) | 91(36.1) |  | 32(53.4) | 105(36.4) |  |
| Bolsa Família cash transfer program |  |  |  | 0.04 |  |  | 0.23 |
| No | 275(79.0) | 69(71.9) | 206(81.7) |  | 44(73.3) | 231(80.2) |  |
| Yes | 73(21.0) | 27(28.1) | 46(18.3) |  | 16(26.7) | 57(19.8) |  |
| Mother age, y (N=345) |  |  |  | 0.17 |  |  | 0.74 |
| <20 | 43(12.8) | 9(10.2) | 34(13.8) |  | 6(10.5) | 37(13.3) |  |
| 20-29 | 174(51.9) | 41(46.6) | 133(53.8) |  | 32(56.2) | 142(51.1) |  |
| ≥30 | 118(35.3) | 38(43.2) | 80(32.4) |  | 19(33.3) | 99(35.6) |  |
| Maternal education, y (N=345) |  |  |  | 0.13 |  |  | 0.38 |
| <8 | 77(22.4) | 26(27.9) | 51(20.3) |  | 16(26.7) | 61(21.5) |  |
| ≥8 | 267(77.6) | 67(72.1) | 200(79.7) |  | 44(73.3) | 223(78.5) |  |
| Paternal education, y (N=345) |  |  |  | 0.66 |  |  | 0.40 |
| <8 | 71(24.5) | 20(26.3) | 51(23.8) |  | 14(29.2) | 57(23.5) |  |
| ≥8 | 219(75.5) | 56(73.7) | 163(76.2) |  | 34(70.8) | 185(76.5) |  |
| Maternal occupation |  |  |  | 0.85 |  |  | 0.58 |
| Unpaid | 212(61.5) | 57(60.6) | 155(61.8) |  | 35(58.3) | 177(62.1) |  |
| Paid | 133(38.5) | 37(39.4) | 96(38.2) |  | 25(41.7) | 108(37.9) |  |
| Number of siblings |  |  |  | 0.02 |  |  | 0.28 |
| 1 | 147(42.2) | 36(37.5) | 111(44.1) |  | 21(35.0) | 126(43.7) |  |
| 2 | 106(30.5) | 24(25.0) | 82(32.5) |  | 18(30.0) | 88(30.6) |  |
| ≥3 | 95(27.3) | 36(37.5) | 59(23.4) |  | 21(35.0) | 74(25.7) |  |
| Number of antenatal care visits * |  |  |  | 0.23 |  |  | 0.40 |
| <6 | 33(9.6) | 12(12.6) | 21(8.4) |  | 4(6.67) | 29(10.2) |  |
| ≥6 | 312(90.4) | 83(87.4) | 229(91.6) |  | 56(93.3) | 256(89.8) |  |
| Type of delivery (345) |  |  |  | 0.19 |  |  | 0.41 |
| Vaginal | 163(47.3) | 39(41.5) | 124(49.4) |  | 25(42.4) | 138(48.3) |  |
| Cesarean section | 182(52.7) | 55(58.5) | 127(50.6) |  | 34(57.6) | 148(51.7) |  |
| Gender |  |  |  | 0.05 |  |  | 0.61 |
| Female | 158(45.4) | 44(45.8) | 146(57.9) |  | 29(48.3) | 129(44.8) |  |
| Male | 190(54.6) | 52(54.2) | 106(42.1) |  | 31(51.7) | 159(55.2) |  |
| Child age (months) |  |  |  | 0.05 |  |  | <0.001 |
| <12 | 128(36.9) | 27(28.4) | 101(40.1) |  | 8(13.3) | 120(41.8) |  |
| ≥12 | 219(63.1) | 68(71.6) | 151(59.9) |  | 52(86.7) | 167(58.2) |  |
| Birth weight |  |  |  | 0.10 |  |  | 0.22 |
| <2500g | 42(12.1) | 16(16.7) | 26(10.3) |  | 10(16.7) | 32(11.1) |  |
| ≥2500g | 306(87.9) | 80(83.3) | 226(89.7) |  | 50(83.3) | 256(88.9) |  |
| Hospitalization in the last 12 months (N=345) |  |  |  | 0.48 |  |  | 0.001 |
| No | 305(88.4) | 83(86.5) | 222(89.2) |  | 45(76.3) | 260(90.9) |  |
| Yes | 40(11.6) | 13(13.5) | 27(10.8) |  | 14(23.7) | 26(9.1) |  |
| Breastfeeding |  |  |  | 0.05 |  |  | <0.001 |
| No | 160(45.9) | 52(54.2) | 108(42.8) |  | 41(68.3) | 119(41.3) |  |
| Yes | 188(54.1) | 44(45.8) | 144(57.2) |  | 19(31.7) | 169(58.7) |  |
| Child attend day care (N=345) |  |  |  | 0.18 |  |  | 0.39 |
| No | 46(13.3) | 87(90.6) | 214(85.3) |  | 50(83.3) | 251(87.5) |  |
| Yes | 86.7) | 9(9.4) | 37(14.7) |  | 10(16.7) | 36(12.5) |  |

The majority of children presenting psychomotor and social developmental delays belonged to the most social excluded families, based on results from the chi-square test which showed differences in psychomotor and social developmental delay between the family social groups. Not being enrolled in the “Bolsa Família” cash transfer showed association with psychomotor developmental delay (P-value=0.04). The number of siblings was associated with social development delay (P-value=0.02). Have been hospitalized in the last 12 months and not breastfed were associated with social development delay (P-value <0.001) (Table 1).

Table 2 shows the results from the four hierarchical multiple logistic regression models for the associations between all variables and psychomotor development delay. Children in the most excluded social groups had a greater odds of psychomotor development delay (OR=3.4, 95% CI=1.14; 10.55) compared to those of the least vulnerable groups. Also, children who were hospitalized in the last 12 months had a greater odds of presenting psychomotor development delay (OR= 2.64, 95% C=1.10; 6.31) than children who were not hospitalized in the past 12 months. Children who were breastfed were 0.53 times less likely (95% CI: 0.23; 0.98) to have psychomotor development delay than children who were not breastfed.

**Table 2.**
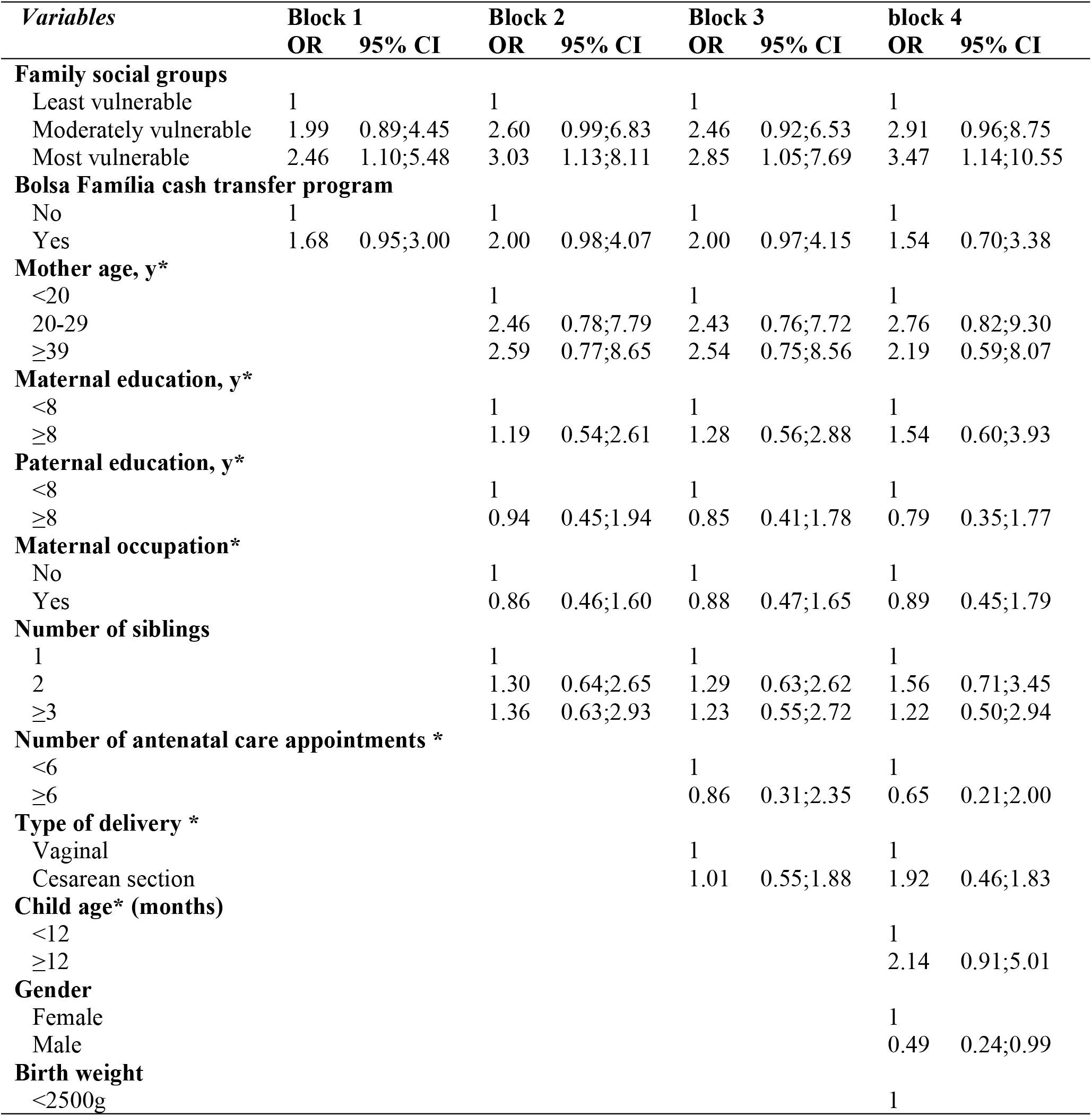
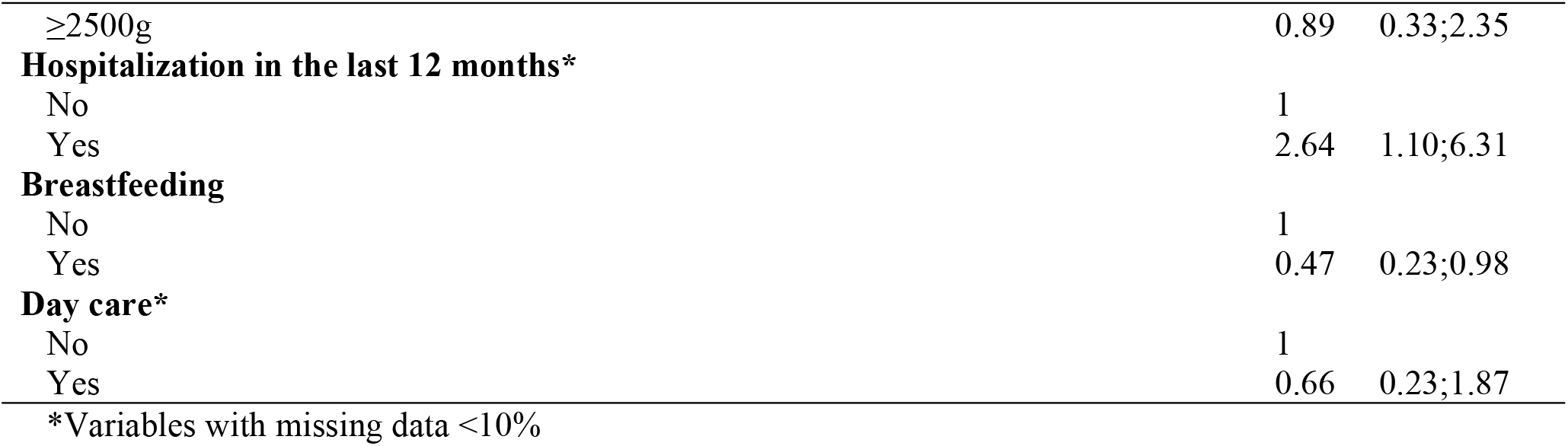
Predictors of psychomotor development delay in children under 3 years old (n = 358). Municipality of the São Paulo State, Brazil, 2013.

Table 3 shows the results from the hierarchical multiple logistic regression for the associations with children’s social development delay. Children in the moderately social excluded group (OR=3.20; 95% CI=1.10; 9.31) and in the most social excluded group had a greater odds of social developmental delay (OR=3.09; 95% CI=1.05; 9.02) comparing those of least social excluded groups. We found no statistical association with the other variables.

**Table 3.**
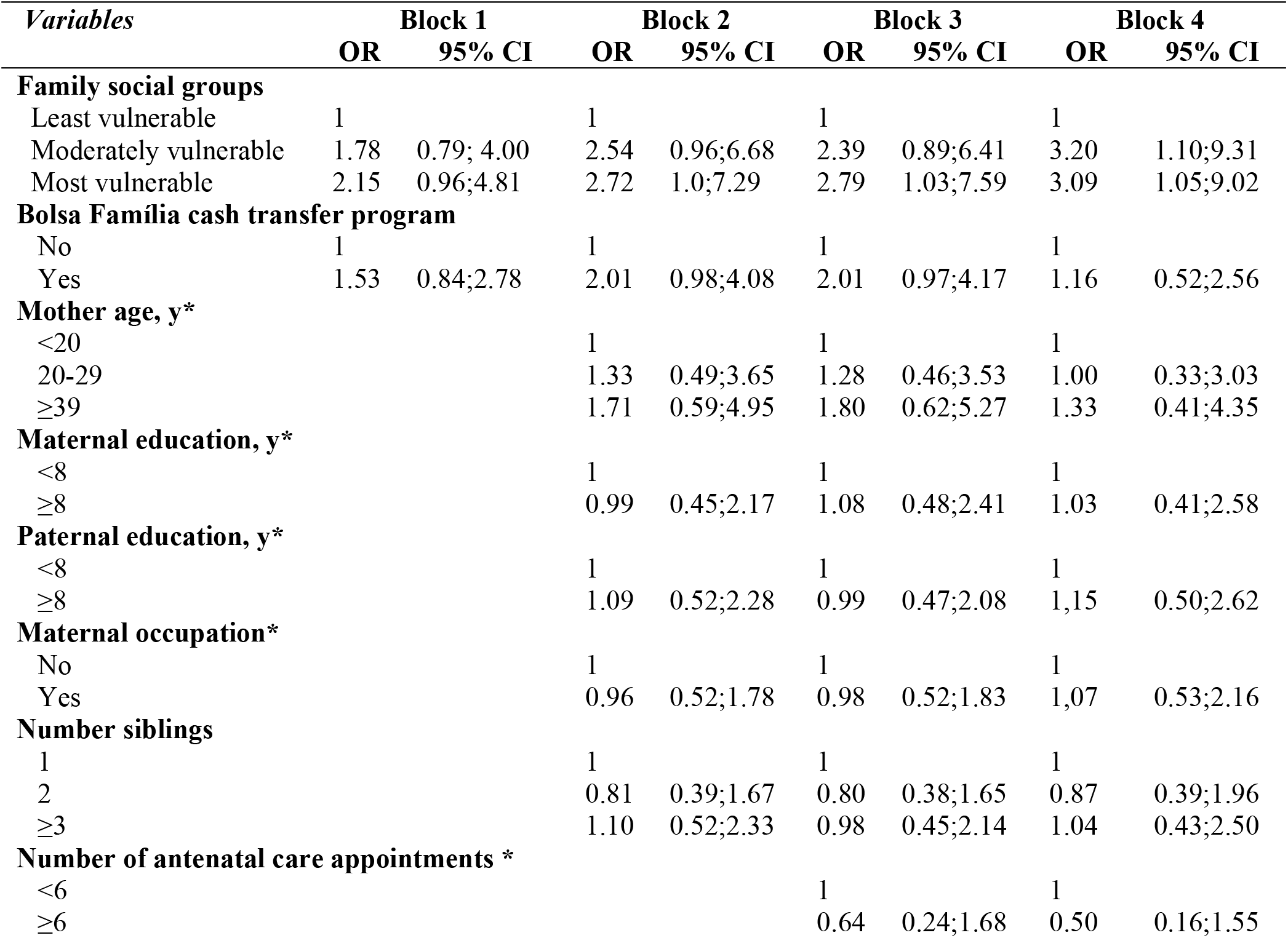
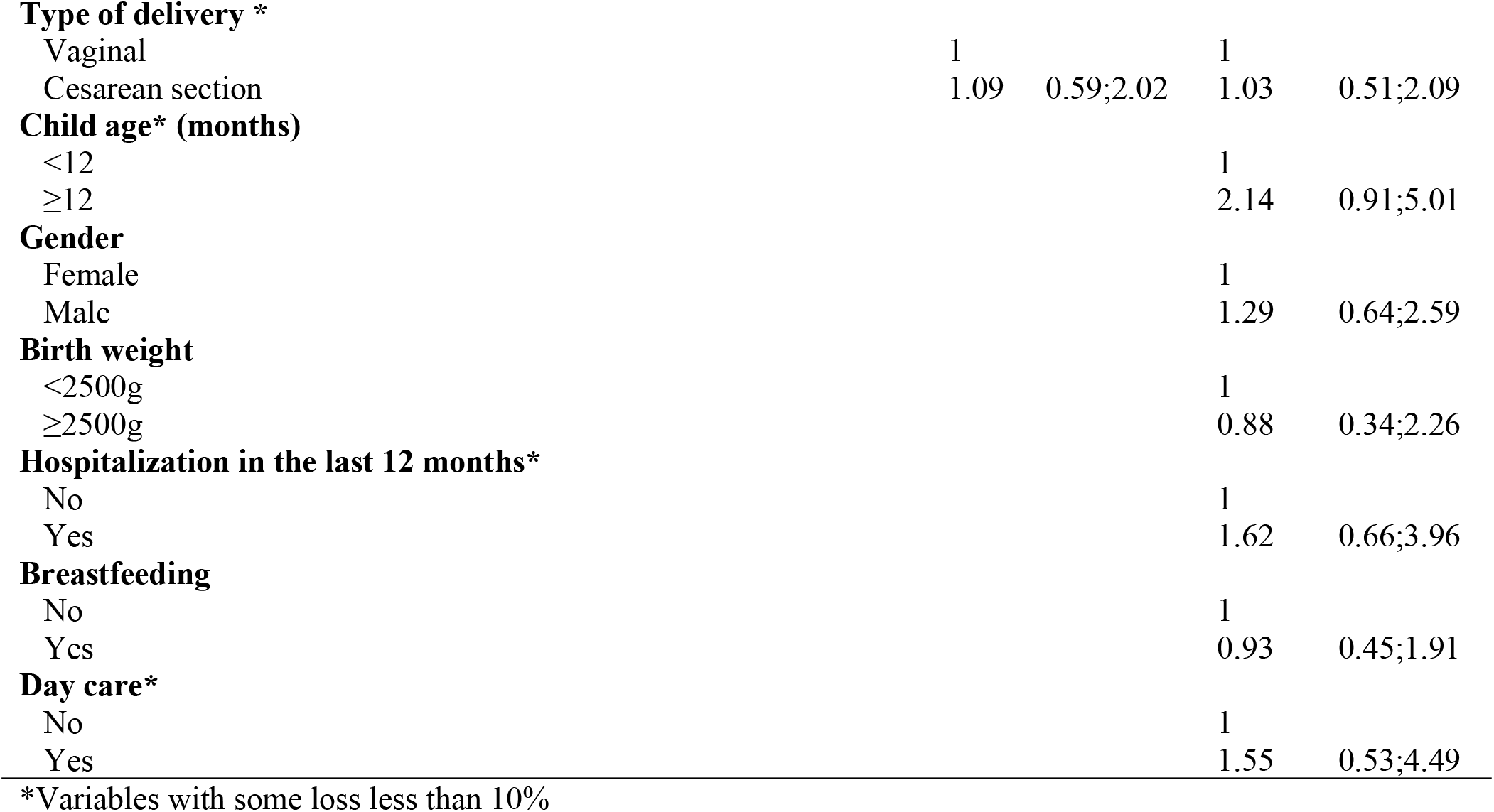
Predictors of social development delay in children between under 3 years old (n = 358). Municipality of São Paulo State, Brazil, 2013.

## Discussion

In the present study, we explored the association between family social exclusion and early child psychomotor and social delay, using a new social inequality index to measure family social exclusion. Our results confirm that children from families that belonged to the most excluded social groups had a greater odds of presenting psychomotor and social delay, which confirms our hypothesis. Previous studies have similarly shown associations between family social determinants and ECD delays using a variety of different measures of social exclusion and child development [19-22].

The majority of prior studies have focused only on socioeconomic characteristics without including variables to measure social exclusion. For instance, a community-based cross-sectional study in South-West Ethiopia [29] observed that children in extreme poverty performed more poorly than the reference children in all the developmental domains comparing with children from the richest group. The authors explained that differences in the developmental outcomes between the two groups might be related because of the adverse early experiences and the socioeconomic inequality of children in extreme poverty [29]. Another study using data from Cambodia, China, Mongolia, Papua New Guinea, Timor-Leste, and Vanuatu found that poor socioeconomic gradients were directly related to lower levels of children’s social-emotional, cognitive and language development and emergent literacy [30]. A comprehensive review also showed that poverty and food insecurity could detrimentally affect the development of young children aged from birth to 3 years [31].

Our findings are also consistent with prior studies in Brazil regarding the social determinants of ECD. Biscegli et al. (2007) found that of the 37% of children with suspected development delay, the majority belonged to families with low income. De Paiva et al. (2010) verified a high prevalence of children with language development delays in families of the lower quartile of the socioeconomic index. Veleda et al. (2011) identified that the highest proportion of children with psychomotor development delay belonged to lower-income families.

These associations can be explained by the fact that the quality of housing, sanitation, hygiene, and food conditions are poorer in socially disadvantaged families, contributing to the increase of diseases and inadequate ECD [6]. Also, a study evaluating child development among employed and unemployed parents revealed the negative influence of parent’s unemployment status on ECD [33]. Indeed, analysis between economics and developmental psychology suggested that job loss can affect mental health academic performance and increase income inequality, particularly among African-American students and those from the poorest families [35].

Our study also confirms another documented correlate of ECD. More specifically, breastfed children had a lower chance of present a delayed psychomotor development compared with children who were not breastfed after controlling for other variables. Breastfeeding is associated with numerous benefits for the developing infant in both the short and the long term, including psychological development [36]. A study that examined how breastfeeding during the first 4 months of life affects the mental and psychomotor development of children aged 6 and 12 years showed that children who were breastfed for at least four months had a higher psychomotor development at 6 and 12 months of age [36]. Breastmilk contains adequate amounts of micronutrient and is vital for optimal growth and development. On the other hand, children who have been hospitalized in the last 12 months had a higher change of delayed psychomotor development. Hospitalization negatively affects the child’s social environment and routine activities and habits, which may impact development due to stressful procedures and experiences [37].

In our study, there was no association between the “Bolsa Família” cash transfer program and child development. However, a recent study showed the importance of “Bolsa Família” as a benefit for low-income families and reducing infant mortality, especially in poorer regions, where the return of the federal health investments in addressing infant mortality is higher [38]. However, other studies have shown the importance of poverty alleviation programs for ECD [9,39]. In Mexico, for example, cash transfer programs have shown significant improvement in height, cognition, and language development [38].

This study advances the understanding of child development from the perspective of social inclusion, as it uses a novel indicator developed originally in Latin America to classify family social exclusion. A limitation of the study is that the ECD assessment instrument used in this study, as available in the Child Health Handbook, is widely used by physicians and nurses working in the primary health care setting in Brazil [27] and has proven useful in tracking early changes in child development, reiterating the importance of its use in primary care. However, it is not specific to diagnose changes in the child’s development and, therefore, constitutes a limitation in this study. Additionally, we did not have information about father and mother behavior as stimulation, interaction, and mental health that could impact child development.

## Conclusion

Inequalities begin during pregnancy and in the early years of life. Children who experience adverse experiences from the beginning of life are at a disadvantage to reach their potential development [6]. In this context, families are one of the most crucial and nurturing environments for children. There is a great need to understand how family inequality influences ECD in different contexts worldwide, especially in Brazil, so this study is of great high significance for filling the gap.

Social exclusion can affect children’s living and learning conditions and their access to nutrition, education, and health. For instance, poorer households may face greater difficulties in providing nutritious food and responsive care for young children. In addition to poverty, social exclusion may limit the availability of educational resources and the utilization of healthcare services for children. Consequently, inequality in socioeconomic resources can result in poor early child development. Furthermore, there is an urgent need to incorporate developmental monitoring into the existing well-child healthcare visits at the primary care level to ensure timely identification and early interventions for children with developmental delays. [40]. Therefore, we suggest that the government and policymakers can provide social and economic support and regular analyses of the families’ social inclusion to ensure that the less privileged women and children are effectively reached with essential interventions.

## Data Availability

https://doi.org/10.7910/DVN/KUDRJI

## Acknowledgments

We are thankful to all participants and professional health workers involved in this study, to the Municipal Health Secretariat, and the Primary Health Care Units.

